# Antenatal depressive symptoms are strongly predicted by the severity of pre-menstrual syndrome: results of partial least squares analysis

**DOI:** 10.1101/2022.06.21.22276727

**Authors:** Yoshiko Abe, Wandee Sirichokchatchawan, Ussanee Sangkomkamhang, Sirina Satthapisit, Junpen Suwimonteerabutr, Michael Maes

## Abstract

**Objective:** Antenatal depression (AD) is the commonest morbidity during pregnancy. There is evidence that premenstrual syndrome (PMS) and AD share common immune-inflammatory pathways. Few studies have assessed the relation between the affective symptoms of PMS and AD. The present study aims to evaluate the association between the severity of depressive PMS and AD in early and late pregnancy.

**Methods:** Women in early pregnancy (<=16 weeks) were recruited and followed until late pregnancy (>=20 weeks). The Premenstrual Symptoms Screening Tool (PSST) was used to assess PMS and AD symptoms were assessed using the Edinburgh Postnatal Depression Scale (EPDS). Results: The PSST score was significantly and positively associated with the EPDS scores both in early and late pregnancy. Up to 57.6% of the variance in the early EPDS score was explained by the regression on the first factor extracted from 10 depression and anxiety PSST items (dubbed the DepAnx PSST domain), PSST item insomnia, relation dissatisfaction, and the Abuse Assessment Screen item 1 (partner abuse). Additionally, 6.3% of the variance in the PSST DepAnx domain was explained by the regression on the mental-physical neglect score of the Adverse Childhood Experiences Questionnaire. There were specific indirect effects of PSST DepAnx (p<0.001), insomnia (p=0.041), relation dissatisfaction (p=0.023) and partner abuse (p=0.007) on the late EPDS which were mediated by the early EPDS score.

**Conclusion:** The affective, but not psychosomatic, symptoms of PMS strongly predict depressive symptoms during pregnancy suggesting that the pathophysiology of affective PMS symptoms overlap with those of AD.

## 1. Introduction

Antenatal depression (AD) is the commonest morbidity during pregnancy with an estimated prevalence of around 18% (Bennett et al., 2004; Gavin et al., 2005). Untreated AD is a global public health concern with adverse sequelae for maternal and offspring well-being (Roomruangwong et al., 2017). AD increases the risk of preterm birth, low birth weight, operational delivery, and preeclampsia, and also affects child neurocognitive development and behavior outcomes later in life (Gentile, 2017; Grigoriadis et al., 2013; Grote et al., 2010; Hu et al., 2015; Jarde et al., 2016). A recent systematic review of large population-based studies found that depression was more common during pregnancy than after delivery (17% and 13%, respectively), and an average of 39% among those who experienced AD went on to have postpartum depression (PPD), which makes AD as one of the major risk factors of PPD (Underwood et al., 2016). Furthermore, PPD is associated with maternal and familiar distress, suicidal risk, and adverse childhood experiences including child abuse (Abe et al., 2022; Ayers et al., 2019; Duan et al., 2019). AD severity is strongly predicted by psychological stressors such as relation dissatisfaction, unintended pregnancy, abuse by the partner and early lifetime trauma (Abe et al., 2022).

Many women of reproductive age also experience one or more emotional, behavioral, and physical symptoms in the premenstrual phase of the menstrual cycle, which is known as premenstrual syndrome (PMS) (Onwude, 2015; Roomruangwong and Maes, 2021). The point prevalence of PMS is estimated between 20-30% (Yonkers and Simoni, 2018). Although most of the symptoms are mild, 1.2-6.4% of reproductive women suffer from moderate to severe symptoms, dubbed premenstrual dysphoric disorder (PMDD), which is defined in the Diagnostic and Statistical Manual of Mental Disorders, Fifth Edition (Yonkers and Simoni, 2018). The typical mood and behavioral symptoms of PMS/PMDD, namely irritability, tension, depressed mood, tearfulness and mood swings, may lead to substantial social, occupational, and interpersonal impairment (Onwude, 2015; Roomruangwong and Maes, 2021). The symptoms often worsen substantially 6 days before the menses and peak around 2 days before the menses whilst there is a symptom-free interval before ovulation (Yonkers et al., 2008).

There is now evidence that PMS or better Menstrual Cycle – Associated Syndrome (MCAS, which offers better diagnostic criteria than PMS and PMDD) (Roomruangwong and Maes, 2021) is characterized by specific alterations in sex hormones indicative of relative corpus luteum insufficiency and alterations in the uterine-brain axis with specific alterations in chemokine levels, bacterial antigens and nitro-oxidative stress biomarkers (Roomruangwong et al., 2019; Roomruangwong et al., 2020a; Roomruangwong et al., 2020b). In addition, other research also indicates that inappropriate inflammatory responses and oxidative stress might contribute to the etiology of PMS (Granda et al., 2021). A recent systematic review concluded that PMS may be accompanied by a decreased antioxidant status and increased inflammatory potential (Granda et al., 2021). Furthermore, menstruation-related symptoms, including low abdominal cramps, gastro-intestinal (GI) and pain symptoms, are strongly associated with inflammatory and antioxidant biomarkers (Roomruangwong et al., 2021). It is important to note that both major depression (Maes and Carvalho, 2018) and PPD (Roomruangwong et al., 2018) are characterized by activated immune-inflammatory and oxidative stress pathways. As such, PMS and AD share common pathways and, therefore, there may be a clinical association between PMS and AD. Nonetheless, few studies have shown that women with PMS symptoms show more severe depressive symptoms throughout pregnancy and increased risk towards antenatal depression (M. Sugawara, 1997; Pataky and Ehlert, 2020).

There is also evidence that adverse childhood experiences (ACEs) are strongly associated with the onset of AD (Abe et al., 2021), major depressive disorder (Maes et al., 2019; Maes et al., 2022), and possibly premenstrual syndrome (PMS) (Jacobs et al., 2015; Yang et al., 2022). Such findings suggest that ACEs may underlie the association between PMS and AD. However, no data have investigated whether ACEs may explain the co-occurrence of the affective or physiosomatic (previously denoted as psychosomatic) symptoms of PMS and depressive symptoms during pregnancy.

Hence, the present study aims to determine the association between PMS severity and its symptomatic subdomains (affective versus physiosomatic) and AD during early and late pregnancy, as well as whether this association can be explained by other psychological stressors, such as ACEs. Based on the shared pathophysiology of PMS and AD, the primary hypothesis is that PMS predicts AD symptoms during early and late pregnancy and that ACEs impact those associations.

## 2. Methods

### 2.1. Participants

This study was conducted at the Khon Kaen Hospital Antenatal Care (ANC) Center in Khon Kaen, Thailand. The hospital is classified as a regional hospital. Khon Kaen is one of the most populous provinces in Thailand’s Northeast. A trained researcher evaluated the eligibility of all pregnant women attending the ANC based on predetermined inclusion and exclusion criteria. Thai pregnant women between the ages of 18 and 49, residing in the Northeast of Thailand, planning to continue ANC treatment in the hospital until birth, being able to read and write Thai, and possessing a mobile phone met the inclusion criteria. Women diagnosed with any axis-I mental disorder other than depression, including schizophrenia, bipolar disorder, generalized anxiety disorder, substance abuse disorders, and psycho-organic disorders, as well as neuroinflammatory or (auto)immune disorders (such as psoriasis, rheumatoid arthritis, inflammatory bowel disease and systemic lupus erythematosus), and women taking antidepressants or mood stabilizers were excluded from the study. All women provided their informed consent in writing. Institutional Review Boards (IRBs) at both Khon Kaen Hospital and Chulalongkorn University (KEF62036 and COA No. 280/2019) approved the study.

### 2.2. Data collection

During normal prenatal appointments, early (gestational week =<16) and late (gestational week >=20) pregnancy data were collected. The sociodemographic data included age, pre-pregnancy body mass index (BMI), level of education, number of children, and age of the partner. Medical records were reviewed for obstetric information such as AD/PPD history, gravidity, parity, and cesarean delivery history. Relationship satisfaction was assessed using a 4-point Likert scale from “satisfied” to “dissatisfied”.

The Edinburgh Postnatal Depression Scale (EPDS) was used to evaluate AD symptoms (Cox et al., 1987). EPDS is the most popular and reliable instrument for assessing depression during pregnancy (Bunevicius et al., 2009). It comprises of 10 questions on a four-point Likert scale (scores range from 0 to 3; total score varies from 0 to 30) and measures the mother’s mood throughout the previous week. The validated cutoff value for screening pregnant women in the Thai version of the EPDS was 10 (area under the curve: AUC 0.84, sensitivity 60%, and specificity 90%) (Pitanupong et al., 2007). This scale has strong reliability and validity, as indicated by a research conducted in the upper Northeast area of Thailand among pregnant women in their third trimester (Item-Objective-congruence: IOC = 0.90; Cronbach’s Alpha = 0.83) (Phoosuwan et al., 2017).

The Thai version of the Premenstrual Symptoms Screening Tool (PSST) was used to assess PMS symptoms. The premenstrual symptoms (#1-14: 14 items) and its consequences in various situations (A-E: 5 items) are measured on a four-point Likert scale ranging from “not at all” to “mild” to “moderate” to “severe” and scored from 1 to 4 (Steiner et al., 2003). Cronbach’s alpha was more than 0.9 for all domains when evaluating the validity of the Thai version (Chayachinda et al., 2008).

The Multidimensional Scale of Perceived Social Support (MSPSS) measures perceived social support (Zimet et al., 1988). The MSPSS is comprised of 12 items scored on a seven-point Likert scale. Family support, friend support, and significant other support are subdomains. Wongpakaran et al. (2018) developed a validated Thai version. In addition, this questionnaire was utilized to evaluate pregnant women’s social support in Thailand’s Northeast area (Phoosuwan et al., 2018).

The Abuse Assessment Screen (AAS) was used to assess intimate relationship abuse (McFarlane, 1992). AAS is one of the most extensively used screening instruments for intimate partner violence in the pregnant population and consists of five yes/no questions (Neha A. Deshpande, 2013). “Yes” item to any question implies abuse. Previously, we detected that AAS item 1 is most important item to predict AD (Abe et al., 2021).

The Adverse Childhood Experiences Questionnaire (ACE questionnaire) was used to evaluate adverse childhood experiences (Felitti et al., 1998). It comprises 28 questions organized into 10 items, namely psychological abuse, physical abuse, sexual abuse, mental neglect, physical neglect, domestic violence, substance misuse in the family, family psychiatric disease, separation or divorce in the family, and family members with criminal behaviors. Using all ACE items, we conducted factor analysis and identified two factors, namely a) mental and physical neglect (dubbed ACEneglect) and b) psychological, physical, and sexual abuse, domestic violence, a family history of substance misuse, and a family history of psychiatric disease (dubbed ACE Family-Dysfunction & Abuse or ACE-FDA) (Abe et al., 2022). Therefore, we used the ACEneglect and ACE-FDA factor scores in the present study.

### 2.3. Data analysis

We utilized analysis of variance (ANOVA) to compare continuous variables between study groups and the Chi-squares (χ2) or Fisher’s exact probability test to compare categories. Using Pearson’s product-moment correlation coefficients, associations between continuous variables were analyzed. We used Generalized Estimating Equation analysis, repeated measures, to examine the differences in EDPS scores between early and late pregnancy. Stepwise (automatic) multiple regression analysis was utilized to delineate the most influential variables predicting the early and late EPDS scores, which were entered as output variables. Outcomes of multiple regression analysis were always checked for multicollinearity and heteroscedasticity. The various subdomains of the PSST items were investigated using principal component analysis. The Kaiser-Meyer-Olkin measure of sampling adequacy (KMO) statistic and Bartlett’s test were utilized to examine the data’s factoriability. The be accepted as an adequate component, the variance explained should be > 50.0% and all items should load highly (>0.6) on the component. All statistical analyses were conducted using version 25 of IBM SPSS for Windows.

In addition, we utilized Partial Least Squares (PLS) analysis to investigate the causal effects of ACEs on the EPDS scores and whether these effects are mediated by PSST scores. The latter were introduced as a latent vector extracted from the PSST items. The AAS1, unplanned pregnancy and relation satisfaction (and socio-demographic data) were entered as other predictors of the EPDS scores. The latter variables, early and late EPDS, and the ACE subdomains were entered as single indicators. Complete PLS analysis was performed when the model quality data met the following predetermined criteria: a) the outer model should have adequate construct validity with AVE>0.5, Cronbach alpha > 0.7, rho_A>0.75 and composite reliability >0.8;. b) the standardized root mean square (SRMR) should be < 0.080 (conservative criterion); c) the original values of the squared Euclidean distance (d ULS) and the geodesic distance (d G) are included in the confidence intervals, d) blindfolding demonstrates adequate cross validated redundancies; and e) PLS Confirmatory Tetrad Analysis shows that the constructs are not mis-specified as a reflective model. Consequently, we conducted complete PLS analysis with 5,000 bootstrap samples and computed pathway coefficients with exact p-value, indicator loadings in the outer model (with p values), specific indirect effects, total indirect effects and total effects.

## 3. Results

### Socio-demographic and clinical data

**Table 1** shows the socio-demographic data of women with and without antenatal depression. There were no significant differences in age, BMI, education, number of children, age of the partner, score of a significant other and family, sexual abuse, mental neglect, and psychiatric family history. There were significant differences in score of friends, total PSS, EPDS total, psychological and physical abuse, physical neglect, domestic violence, substance use disorder (SUD) in family, household dysfunction, and ACE total.

**Table1.** Baseline characteristics of women with and without antenatal depression (AD)

|  |  |  |  |  |  |  |  |
| --- | --- | --- | --- | --- | --- | --- | --- |
| | AD | | No AD | | F/$\chi^2$/FFPT | df | p-value |
|  | n=34 |  | n=86 |  |  |  |  |
| Age (years) | 27.4 | 6.0 | 27.1 | 5.3 | 0.08 | 1/118 | 0.772 |
| Pre-pregnancy BMI (kg/m2) | 23.2 | 3.8 | 23.4 | 5.8 | 0.02 | 1/118 | 0.898 |
| Education level | 3.3 | 1.3 | 3.6 | 1.3 | 1.18 | 1/118 | 0.280 |
| Number of children | 0.7 | 0.7 | 0.7 | 0.6 | 0.32 | 1/118 | 0.575 |
| Partner age (years) | 31.4 | 8.6 | 29.7 | 6.0 | 1.44 | 1/118 | 0.232 |
| MSPSS significant other | 5.4 | 1.0 | 5.6 | 1.2 | 0.83 | 1/118 | 0.365 |
| MSPSS family | 5.9 | 1.0 | 6.2 | 0.8 | 3.35 | 1/118 | 0.070 |
| MSPSS friends | 4.7 | 1.3 | 5.6 | 1.0 | 13.48 | 1/118 | <0.001 |
| MSPSS total | 5.4 | 0.9 | 5.8 | 0.8 | 7.14 | 1/118 | 0.009 |
| Total PSST | 32.9 | 6.4 | 24.0 | 4.8 | 67.70 | 1/118 | <0.001 |
| EPDS total | 12.3 | 2.8 | 3.5 | 2.6 | 274.13 | 1/118 | <0.001 |
| Psychological abuse | 3.1 | 1.7 | 2.3 | 0.8 | 13.06 | 1/118 | <0.001 |
| Physical abuse | 2.6 | 1.4 | 2.0 | 0.2 | 12.07 | 1/118 | <0.001 |
| Sexual abuse | 0.1 | 0.3 | 0.1 | 0.4 | 0.39 | 1/118 | 0.536 |
| Mental neglect | 10.9 | 5.0 | 9.2 | 4.1 | 3.68 | 1/118 | 0.057 |
| Physical neglect | 9.5 | 3.7 | 7.7 | 2.9 | 7.27 | 1/118 | 0.008 |
| Domestic violence | 4.6 | 2.0 | 4.1 | 0.4 | 4.84 | 1/118 | 0.030 |
| Substance abuse in the family | 0.2 | 0.5 | 0.0 | 0.2 | 5.86 | 1/118 | 0.017 |
| Family psychiatric illness | 0.1 | 0.3 | 0.0 | 0.2 | 0.19 | 1/118 | 0.666 |
| Separation or divorce in the family | 0.5 | 0.5 | 0.3 | 0.5 | 3.68 | 1/118 | 0.057 |
| Family members are criminals | 0.1 | 0.3 | 0.1 | 0.2 | 0.35 | 1/118 | 0.555 |
| Household dysfunction | 5.4 | 2.7 | 4.5 | 0.8 | 7.73 | 1/118 | 0.006 |
| ACEneglect | 20.4 | 8.0 | 16.9 | 6.3 | 6.17 | 1/118 | 0.014 |
| ACE-FDA | 10.5 | 4.8 | 8.5 | 1.1 | 13.83 | 1/118 | <0.001 |
| ACE total score | 31.6 | 11.1 | 25.9 | 6.8 | 11.81 | 1/118 | <0.001 |
| AD/PPD history (n,%) | 33 | 97.1 | 83 | 96.5 | – | – | 1.000 |
|  | 1 | 2.9 | 3 | 3.5 |  |  |  |
| Planned pregnancy (n, %) | 11 | 32.4 | 65 | 75.6 | 19.61 | 1 | <0.001 |
|  | 23 | 67.6 | 21 | 24.4 |  |  |  |
| Gravidity (times) (n,%) | 13 | 38.2 | 31 | 36.0 | 3.02 | 2 | 0.221 |
|  | 10 | 29.4 | 38 | 44.2 |  |  |  |
|  | 11 | 32.4 | 17 | 19.8 |  |  |  |
| Parity (times) (n,%) | 15 | 44.1 | 34 | 39.5 | 1.79 | 2 | 0.408 |
|  | 14 | 41.2 | 45 | 52.3 |  |  |  |
|  | 5 | 14.7 | 7 | 8.1 |  |  |  |
| History of cesarean delivery (n,%) | 27 | 79.4 | 74 | 86.0 | 0.81 | 1 | 0.410 |
|  | 7 | 20.6 | 12 | 14.0 |  |  |  |
| Relation satisfaction with a partner and family (n,%) | 27 | 79.4 | 83 | 96.5 | – | – | 0.005 |
|  | 7 | 20.6 | 3 | 3.5 |  |  |  |
| AAS1 (n,%) | 23 | 67.6 | 79 | 91.9 | 11.20 | 1 | <0.001 |
|  | 11 | 32.4 | 7 | 8.1 |  |  |  |

F/ $\chi^2$ /FEPT: all results of analysis of variance (ANOVA), analysis of contingency tables ( $\chi^2$ Test) or Fisher's Exact Probability test (FEPT).
Results are shown as mean ± standard deviation, or as ratios with percentage
MSPSS: Multidimensional Scale of Perceived Social Support.
PSST: Premenstrual Symptoms Screening
EPDS: Edinburgh Postnatal Depression Scale.
ACE: Adverse Childhood Experiences.
ACEneglect: Mental and physical neglect
ACE-FDA: Family dysfunction and abuse
AD: Antenatal Depression

### Results of factor analysis

We examined whether the total sum of the PSS is an adequate method to reflect severity of PSS and, therefore, examined whether one latent vector with adequate construct validity could be extracted from the items. The first factor extracted from the 14 PSS symptoms explained only 40.9% of the variance (KMO=0.850), while 3 items showed loading <0.6, namely PSS10, PSS11, PSS12 and PSS14. Consequently, we have examined whether one factor with adequate construct validity could be extracted from the ten remaining PSS items. **Table 2** shows that one factor could be extracted from PSS items 1, 2, 3, 4, 5, 6, 7, 8, 9 and 13 which explained 50.4% of the variance (KMO=0.878), whilst all loadings were > 0.6. We were also able to extract one factor which explained 50.4% of the variance in PSS10, PSS12 and PSS14, while PSS11 did not load highly on this factor. Therefore, we used the first factor extracted from those 10 PSST (labeled PSST depression-anxiety PC) and 3 PSST (dubbed PSST physiosomatic PC) items as well as PSST item insomnia in subsequent analyses. Table 2 also shows that one adequate factor (dubbed PSST interference PC) could be extracted from all five PSS interference data (A, B, C, D and E) which explained 69.3% of the variance and showed high loadings on the five items (>0.716).

**Table2.** Results of principal component analysis of the Premenstrual Symptoms Screening Tool (PSST) items

|  |  | PSST |  |  |  |
| --- | --- | --- | --- | --- | --- |
| Variables | PC | Variables | PC | Variables | PC |
| PSST 10 | 0.641 | PSST 1 | 0.609 | PSST.A | 0.716 |
| PSST 12 | 0.800 | PSST 2 | 0.746 | PSST.B | 0.864 |
| PSST 14 | 0.785 | PSST 3 | 0.722 | PSST.C | 0.816 |
|  |  | PSST 4 | 0.724 | PSST.D | 0.893 |
|  |  | PSST 5 | 0.737 | PSST.E | 0.854 |
|  |  | PSST 6 | 0.750 |  |  |
|  |  | PSST 7 | 0.684 |  |  |
|  |  | PSST 8 | 0.746 |  |  |
|  |  | PSST 9 | 0.729 |  |  |
|  |  | PSST 13 | 0.637 |  |  |
| Component | PC_3PSST | Component | PC_10PSST | Component | PC_interference |
| KMO | 0.61 | KMO | 0.88 | KMO | 0.84 |
| Bartlett's test |  | Bartlett's test |  | Bartlett's test |  |
| $\chi^2$ | 38.85 | $\chi^2$ | 554.04 | $\chi^2$ | 334.24 |
| df | 3 | df | 45 | df | 10 |
| p-value | <0.001 | p-value | <0.001 | p-value | <0.001 |
| Variance explained | 55.6 | Variance explained | 50.4 | Variance explained | 69.0 |

KMO: Kaiser-Meyer-Olkin measure of sampling adequacy

### Differences in PSS scores between women with and without perinatal depression

**Table 3** shows the differences in all 14 PSS items between women with and without perinatal depression. All 14 PSS items as well as all 5 PSS interference scores were significantly higher in women with perinatal depression than in those without. The scores of the total summed PSS, PSST depression-anxiety, PSST physiosomatic and PSST interference PC scores were significantly higher in women with perinatal depression than in those without.

**Table3.** Associations between Premenstrual Symptoms Screening (PSS) score and Antenatal Depression (AD)

| PSST scores | AD<br>n=34 |  | No AD<br>n=86 |  | F | df | p-value |
| --- | --- | --- | --- | --- | --- | --- | --- |
|  | mean | (SD) | mean | (SD) |  |  |  |
| Total PSST | 32.9 | 6.4 | 24.0 | 4.8 | 67.74 | 1/118 | <0.001 |
| PSST1 | 2.8 | 0.6 | 2.3 | 0.7 | 18.06 | 1/118 | <0.001 |
| PSST2 | 2.4 | 0.8 | 1.7 | 0.7 | 26.93 | 1/118 | <0.001 |
| PSST3 | 2.5 | 1.0 | 1.8 | 0.8 | 19.01 | 1/118 | <0.001 |
| PSST4 | 2.0 | 0.8 | 1.1 | 0.3 | 82.97 | 1/118 | <0.001 |
| PSST5 | 2.1 | 0.8 | 1.4 | 0.6 | 26.75 | 1/118 | <0.001 |
| PSST6 | 1.9 | 0.8 | 1.3 | 0.6 | 18.28 | 1/118 | <0.001 |
| PSST7 | 2.0 | 0.7 | 1.4 | 0.6 | 25.39 | 1/118 | <0.001 |
| PSST8 | 2.0 | 0.7 | 0.0 | 0.0 | 31.99 | 1/118 | <0.001 |
| PSST9 | 2.8 | 0.7 | 2.2 | 0.7 | 18.33 | 1/118 | <0.001 |
| PSST10 | 2.6 | 0.8 | 2.1 | 0.9 | 7.62 | 1/118 | 0.007 |
| PSST11 | 2.0 | 0.8 | 1.6 | 0.7 | 9.01 | 1/118 | 0.003 |
| PSST12 | 2.8 | 1.0 | 2.3 | 0.9 | 9.72 | 1/118 | 0.002 |
| PSST13 | 1.9 | 0.8 | 1.3 | 0.5 | 24.43 | 1/118 | <0.001 |
| PSST14 | 3.0 | 0.7 | 2.4 | 0.8 | 14.92 | 1/118 | <0.001 |
| PSST.A | 0.0 | 0.7 | 0.0 | 0.7 | 9.76 | 1/118 | 0.002 |
| PSST.B | 1.8 | 0.8 | 1.3 | 0.6 | 12.26 | 1/118 | <0.001 |
| PSST.C | 1.8 | 0.8 | 1.4 | 0.7 | 8.34 | 1/118 | 0.005 |
| PSST.D | 1.9 | 0.8 | 1.3 | 0.6 | 20.52 | 1/118 | <0.001 |
| PSST.E | 2.3 | 0.9 | 1.4 | 0.7 | 31.40 | 1/118 | <0.001 |
| PSST depression-anxiety | 1.0 | 1.0 | -0.4 | 0.7 | 66.95 | 1/118 | <0.001 |
| PSST physiosomatic | 0.6 | 0.9 | -0.2 | 1.0 | 20.51 | 1/118 | <0.001 |
| PSST interference | 0.6 | 1.0 | -0.3 | 0.9 | 23.55 | 1/118 | <0.001 |

F: all results of analysis of variance (ANOVA)
Results are shown as mean with standard deviation (SD)

### Associations between PSS, EPDS and ACE scores

**Table 4** shows the intercorrelation matrix among PSST scores, EPDS and ACE scores. We found that the EPDS scores in early and late pregnancy were significantly associated with the total PSST score, PSST depression-anxiety PC, PSST interference PC, and PSST insomnia, whilst the early EPDS was additionally associated with the PSST physiosomatic score. The ACEneglect score was positively and significantly associated with the PTSS depression-anxiety PC score. Both EPDS scores were significantly associated with ACEneglect and ACE-FDA scores.

**Table4.** Intercorrelation matrix among Premenstrual Symptoms Screening Tool (PSST), Edinburgh Postnatal Depression Scale (EPDS), and Adverse Childhood Experiences (ACE) scores

|  | EPDS late<br>(n=94) | EPDS early<br>(n=120) | ACEneglect<br>(n=120) | ACE-FDA<br>(n=120) |
| --- | --- | --- | --- | --- |
| EPDS early | 0.510 (<0.001) |  |  |  |
| ACEneglect | 0.360 (<0.001) | 0.291 (0.001) |  |  |
| ACE-FDA | 0.306 (0.003) | 0.310 (<0.001) | 0.350 (<0.001) |  |
| Total PSST | 0.421 (<0.001) | 0.664 (<0.001) | 0.154 (0.092) | 0.131 (0.153) |
| PSST depression-anxiety PC | 0.453 (<0.001) | 0.650 (<0.001) | 0.249 (0.006) | 0.162 (0.077) |
| PSST physiosomatic<br>PC | 0.124 (0.234) | 0.386 (<0.001) | -0.118 (0.199) | 0.026 (0.776) |
| PSST insomnia | 0.261 (0.011) | 0.428 (<0.001) | 0.009 (0.920) | 0.002 (0.984) |
| PSST interference PC | 0.264 (0.010) | 0.469 (<0.001) | 0.179 (0.010) | 0.156 (0.089) |

ACE neglect: Mental and physical neglect
ACE-FDA: Family dysfunction and abuse

### Best predictions of the EPDS scores

**Table 5** shows the results of multiple regression analysis with EPDS as dependent variables and PSST, ACEneglect, ACE-FDA and AAS1 scores (and socio-demographic data including education, body mass index, parity) as explanatory variables. We found that 31.6% of the variance in the endpoint EPDS score was explained by PSST depression-anxiety PC, ACE total, and AAS1 (all positively associated). After entering the baseline EPDS score in the same regression analysis we found that 32.2% of the variance in the endpoint EPDS score was explained by the regression on baseline EPDS and ACE neglect (both positively associated). The third regression shows that 63.1% of the variance in the early EPDS score was explained by the combined effects of PSST depression-anxiety PC, PSST insomnia, ACE1, AAS1, and relation satisfaction. **Figure 1** shows the partial regression of the early EPDS score on the PSST depression-anxiety PC score.

**Table5.**
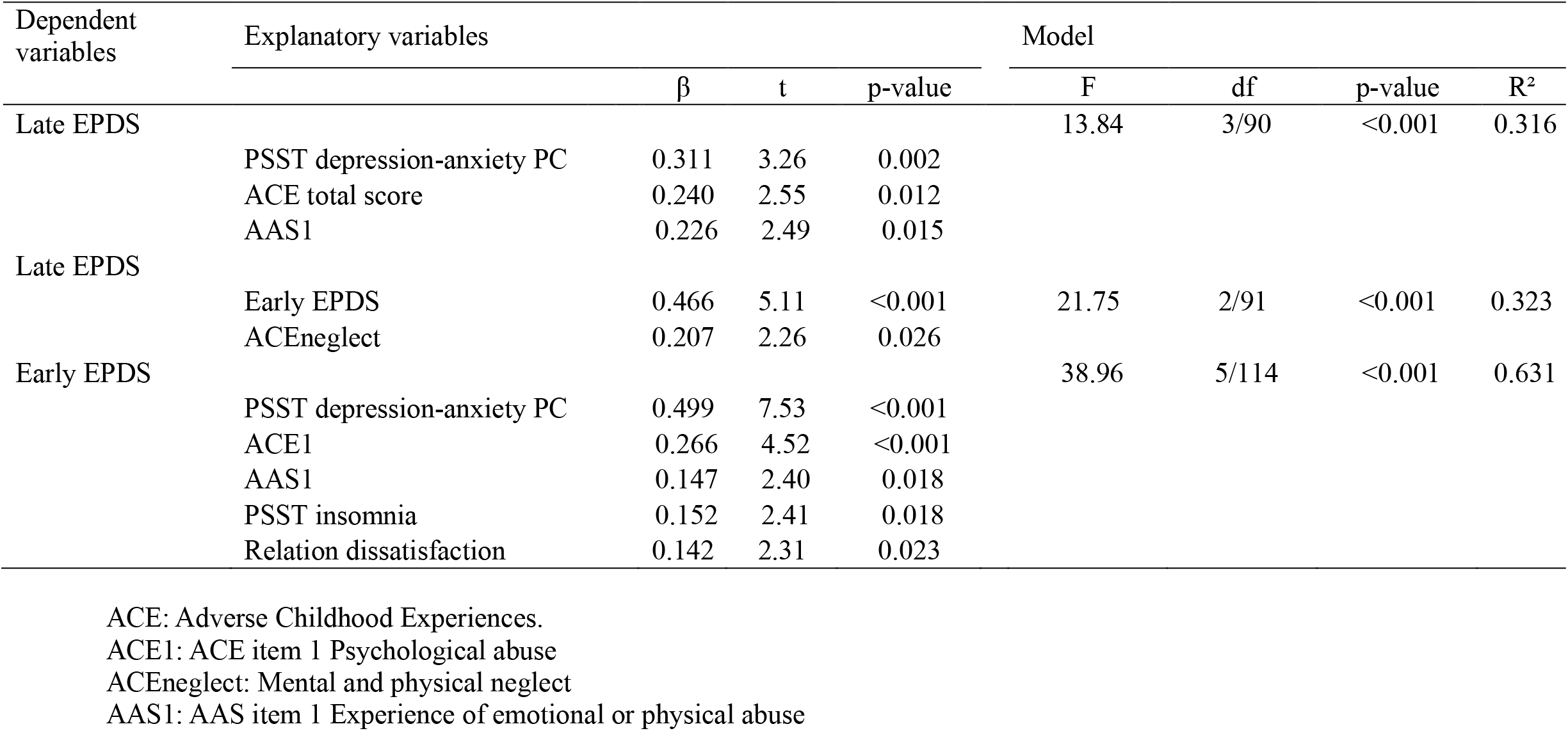
Results of multiple regression analysis with the early and late Edinburgh Postnatal Depression Scale (EPDS) scores as dependent variables

**Figure 1.**
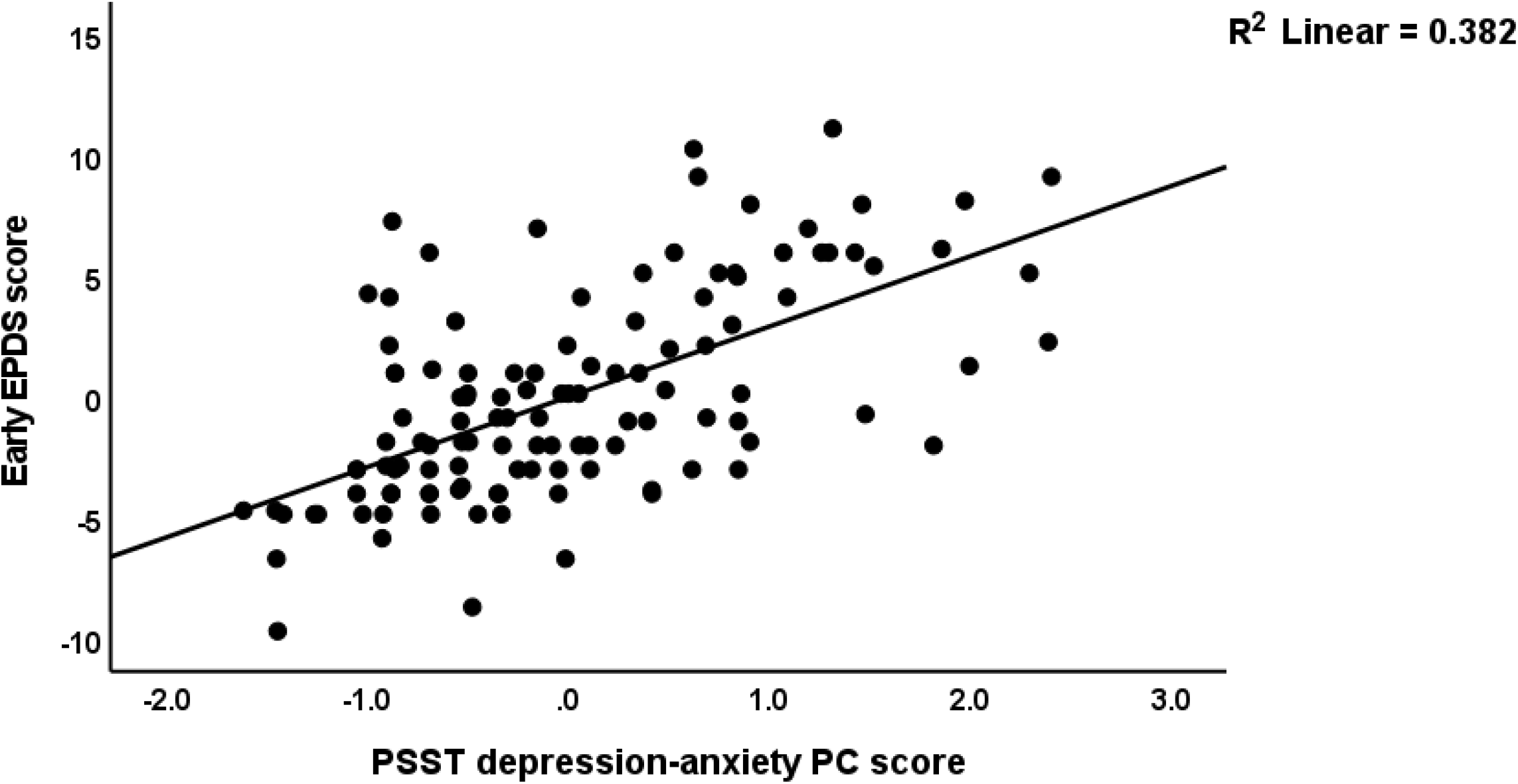
Partial regression of the early Edinburgh Postnatal Depression Scale (EPDS) score on the Premenstrual Symptoms Screening Tool (PPST) depression-anxiety score

**Table 6** shows the results of a GEE repeated measurement analysis with the two EPDS measurements as dependent variables and AAS1 (repeated measures), antenatal depression, time, PSST, ACE scores and items of the ACE as explanatory variables. We found that the changes over time in the EPDS score were best predicted by antenatal depression, time, diagnosis x time interaction, PSST depression-anxiety PC, PSST insomnia, AAS1, ACE1 and ACE neglect. This table also shows the mean EPDS values obtained in early and late pregnancy both in women with and without antenatal depression. The late EPDS score were significantly lower than the baseline EPDS scores while the diagnosis x time interaction pattern may be explained by our findings that in women without antenatal depression there was no effect of time, whereas in those with antenatal depression, the EPDS scores were lower at endpoint than at baseline.

**Table6.** Results of GEE repeated measures analysis with the Edinburgh Postnatal Depression Scale (EPDS) score as dependent variable

| Effects | Early | Late | Wald (df=1) | p-value |
| --- | --- | --- | --- | --- |
| No AD mean (SD) | 4.2 (0.3) | 5.0 (0.4) |  |  |
| AD mean (SD) | 10.4 (0.4) | 5.5 (0.6) |  |  |
| AD |  |  | 38.71 | <0.001 |
| time |  |  | 29.03 | <0.001 |
| time×AD |  |  | 57.93 | <0.001 |
| PSST depression-anxiety PC |  |  | 11.02 | <0.001 |
| PSST insomnia |  |  | 10.45 | 0.001 |
| AAS1 |  |  | 4.51 | 0.034 |
| ACE1 |  |  | 7.74 | 0.005 |
| ACE PC1 |  |  | 5.25 | 0.022 |

AD: Antenatal Depression
AAS1: AAS item 1 Experience of emotional or physical abuse
ACE1: ACE item 1 Psychological abuse
ACE PC1: Mental and physical neglect

### Results of PLS analysis

Figure 2. shows the results of the final PLS model. The quality of the model was adequate with SRMR=0.054 and the outer model of the latent vector extracted from the 10 PSST depression-anxiety items showed accurate construct validity with composite reliability of 0.910, rho_A of 0.897 and Cronbach alpha of 0.890, and AVE=0.503. The outer loadings were all greater than 0.6 at p<0.0001. PLS blindfolding showed an adequate construct crossvalidated redundancy (0.029) and PLS predict showed that this latent vector was not mis-specified as a reflective model. Figure 2 shows the pathway coefficient with exact p values as well as the variance explained. Thus, we found that 25.1% of the variance in late EPDS was explained by early EPDS and ACEneglect scores. Up to 57.6% of the variance in early EPDS was explained by the regression on PSST depression-anxiety PC, PSST insomnia, relation dissatisfaction and AAS1. Finally, 6.3% of the variance in the first latent vector extracted from the PSST depression-anxiety PC was explained by the mental-physical subdomain of the ACE. There were specific indirect effects of the PSST depression-anxiety latent vector (t=4.16, p<0.001), PSST insomnia (t=1.74, p=0.041), relation dissatisfaction (t=1.99, p=0.023) and AAS1 (t=2.47, p=0.007) on the late EDPS score which were all mediated via the early EPDS score. There was a significant indirect effect of the ACEneglect score on late EPDS which was mediated by the path from PSST depression-anxiety PC to early EPDS (t=1.99, p=0.024).

**Figure 2.**
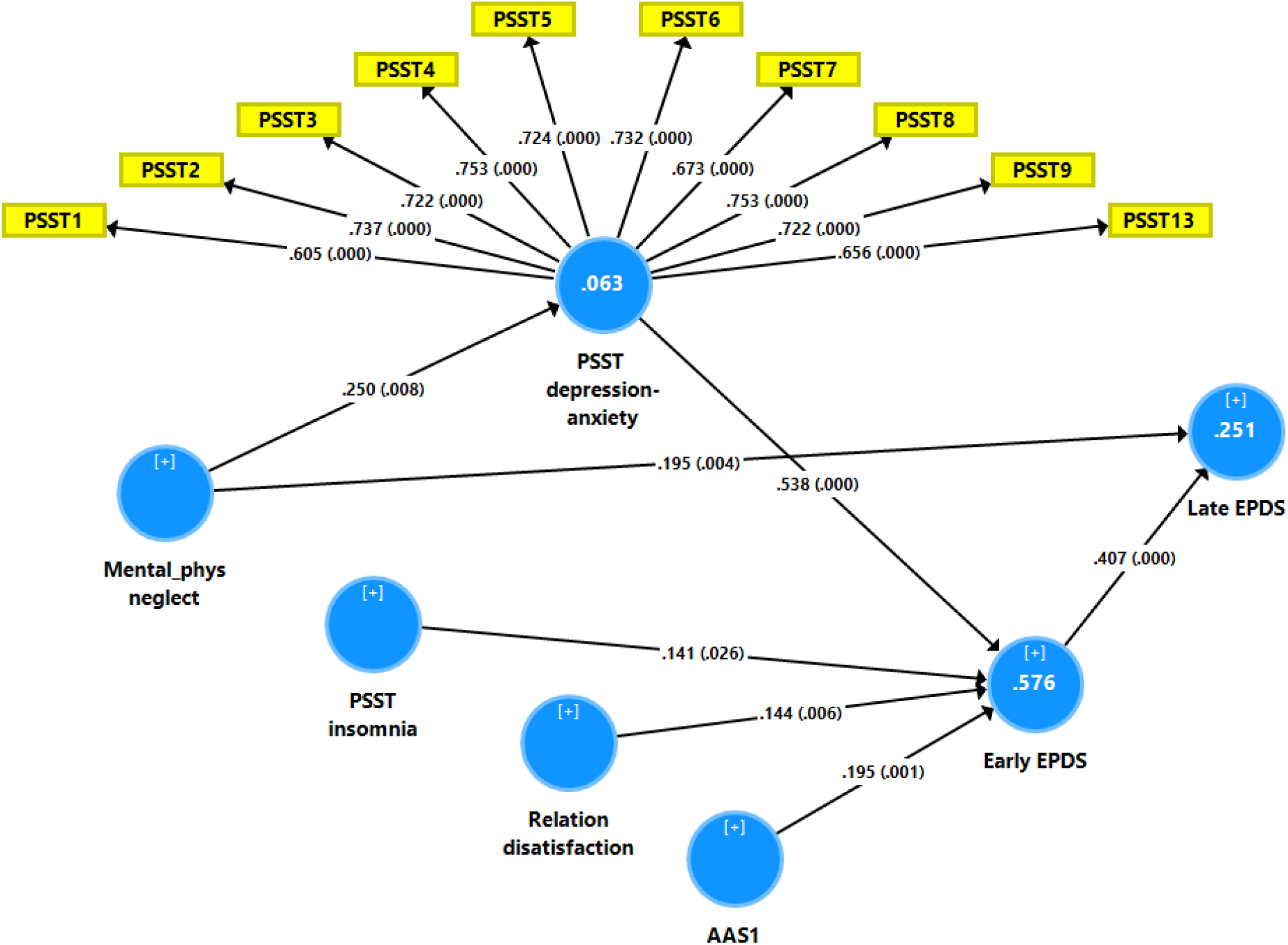
Results of Partial least Squares (PLS) analysis with the early and late Edinburgh Postnatal Depression Scale (EPDS) scores as output variables and the Premenstrual Symptoms Screening Tool (PPST) depression-anxiety and insomnia scores, relation dissatisfaction, and the Abuse Assessment Screen, item 1 (AAS1) score as direct input variables. Mental-physical (mental-phys) neglect predicted the PSST and EPDS scores. The PSST depression-anxiety score was entered as a latent vector extracted from 10 PSST items, whilst all other variables were entered as single indicators. This final PLS model shows only the significant paths. In this model, the PSST depression-anxiety partly mediates the effect of mental-physical neglect on the early and thus late EPDS scores. Shown are path coefficients (with exact p values) and loadings (with p values) on the PSST latent vector. The white figures in the blue circles indicate explained variance.

## 4. Discussion

### Affective and physiosomatic subdomains of the PSST

The first major finding of this study is that the PSST rating scale, when used in pregnant females, does not have sufficient convergence and construct validity. For the sum of all items of a rating scale score to be regarded accurate, the test’s factorial structure must be unidimensional, namely it should contain one general factor explaining more than 50 percent of the variance and all items included should have significant and high loadings (>0.6) on this factor (Hemrungrojn et al., 2022). Without a general factor, the overall sum score will not reflect the overarching construct (Sala et al., 2020). In the present study we found that the first factor extracted from the 14 PSST symptoms explained only 40.9% of the variance while some items showed insufficient loadings of <0.6. We found that the PSST comprises 2 meaningful factors, namely a first with PSST items 1, 2, 3, 4, 5, 6, 7, 8, 9 and 13 (dubbed the PSST depression-anxiety factor) and a second with 3 physiosomatic items (PSS10, PSS12 and PSS14, dubbed the PSST physiosomatic factor) and one solitary item, namely PSST insomnia. One factor with adequate construct validity could be extracted from the five PSST interference data (A, B, C, D and E, dubbed PSST inference PC) and, consequently, we used these two PSST construct scores, insomnia and PSST inference PC in our analysis. In another exploratory factor analysis study, four domains were retrieved namely decreased interest in daily activities, interference with normal function, instability of mood and psychophysical distress (Pacitti et al., 2021).

### Association between PSST scores and prenatal depression

The second major finding of the present study is that all PTSS item and subdomain scores were significantly higher in women with AD than in those without and that the severity of the PSST depression-anxiety factor score, PSST insomnia as well as relation dissatisfaction and the experience of partner abuse significantly predicted depressive EPDS symptoms during pregnancy. In fact, those explanatory variables explained 63.0% of the variance in early pregnancy EPDS scores, whilst the PSST depression-anxiety PC was the single best predictor of severity of AD symptoms. Moreover, the association between the PSST depression-anxiety factor score and depressive symptoms in late pregnancy was mediated by the depressive symptoms in early pregnancy. As such the affective, but not physiosomatic, symptoms of PMS largely predict the depressive symptoms during early and late pregnancy, whereas insomnia has a significant effect be it with a smaller effect size.

Limited studies showed positive associations between PMS and affective disorders during pregnancy. One study identified that women with mood changes (irritability) during the premenstrual period had significantly higher depression scores than those without throughout pregnancy (early, middle, and late pregnancy) and the postpartum period as well (5 days, 1 month, and 6 months after childbirth) (M. Sugawara, 1997). Another study revealed that having symptoms of PMS prior to the current pregnancy was associated with increased odds of antenatal depression (OR: 2.828 [95% CI 1.337–5.983]) (Pataky and Ehlert, 2020). Our findings extend these previous studies showing that PMS is strongly associated with depressive symptoms during pregnancy and, additionally, emphasized the effect of affective symptoms and insomnia, but not physiosomatic, symptoms, during the premenstrual period on depressive symptoms during pregnancy.

### Effects of ACEs

The third main finding of the current study is that mental and physical neglect had a substantial impact on the early pregnancy EPDS score, as well as the late EPDS score. As a result, our findings extend those of a recent meta-analysis reporting that ACEs significantly increase the risk of mother depression and/or anxiety throughout the perinatal period (Racine et al., 2021). Other studies also support that ACEs such as physical, emotional and sexual abuse, physical and emotional neglect, and home dysfunctions) were linked to mental distress during pregnancy (Li et al., 2017; Racine et al., 2018; Shamblaw et al., 2019).

Another significant finding of the current study is that the link between mental and physical neglect and depressed symptoms in early and late pregnancy was partially mediated by the PSST depression-anxiety PC score. Furthermore, the path from PSST depression-anxiety symptoms to early pregnancy EPDS partially mediated the effects of mental and physical neglect on late pregnancy EDPS scores. This is an example of partial mediation whereby the affective symptoms of PMS partially mediate the effects of ACEs on depressive symptoms during pregnancy. This is also an example of distal mediation because the effect of the affective symptoms of PMS on the EDPS score was much more important than the effect of mental and physical neglect on PMS.

A population-based study that looked at the relationship between cumulative and individual ACE exposure and premenstrual disorders, such as PMS and PMDD, discovered a positive, linear relationship between the cumulative number of ACEs and likely premenstrual disorders (PR: 1.12, 95 percent CI: 1.11–1.13). The latter were linked to sexual abuse, emotional neglect, familial violence, a household member’s mental illness, as well as peer and communal violence (Yang et al., 2022). Another study found that women who were abused as children had a significantly higher incidence of premenstrual syndrome (OR: 1.47, 95 percent CI: 1.20–1.88). Physical or emotional maltreatment as a youngster was found to be strongly linked to PMS (Ito et al., 2021). ACEs are risk factors for antenatal mental health disorders, according to a previous mediation analysis, and these effects are mediated by past depressions and unfavorable adult experiences (Lydsdottir et al., 2019). In women with low levels of current resilience, ACE exposure strongly predicts poor mental and behavioral health during pregnancy (Young-Wolff et al., 2019).

### Adverse outcome pathways linking mental and physical neglect, PMS and prenatal depression

The interrelationships among the PSST affective symptoms of PMS and perinatal depressive symptoms and the effects of ACEs may be explained by the effects of adverse outcome pathways. Thus, there is evidence that a) PMS (or better MCAS) and menstrual distress are in part due to immune-inflammatory processes (Roomruangwong and Maes, 2021); b) major depression and perinatal depression are associated with activated immune-inflammatory pathways; and c) ACEs induce an immune-inflammatory and growth factor response which is a major determinant of the phenome of depression (Maes et al., 2022). Moreover, psychosocial stressors including relation dissatisfaction and abuse by the partner during pregnancy may further exacerbate those immune-inflammatory pathways. It is indeed well-known that even mild psychological stressors induce and immune-inflammatory response as well as T cell activation (Maes et al., 1998; Maes et al., 1999). It is interesting to note that these stress effects on the cytokine network are pronounced in subjects with lowered levels of omega-3 polyunsaturated fatty acids (Maes et al., 2000) which is known to predict depressive symptoms in the perinatal period (De Vriese et al., 2003). Consequently, the main hypothesis is that ACE-induced immune responsivity contributes to PMS and perinatal depression, and that the sex-hormone alterations leading to PMS (Roomruangwong and Maes, 2021) aggravate the immune response thereby aggravating perinatal depression.

### Limitations

There are a few limitations to this research. First, the EPDS score was collected by self-report. The PSST and ACEs questionnaires are completed retrospectively by the participants and, hence, they may have been prone to recollection bias. Lastly, our data were obtained at a regional hospital, which is typical of the Northeast area of Thailand and, therefore, our findings may not be generalizable.

## Conclusions

The present study demonstrated that the affective symptoms and sleep disturbance of PMS strongly predict depressive symptoms during early pregnancy, and that this the latter mediates the effects of PMS affective symptoms on late pregnancy depressive symptoms. Moreover, mental and physical neglect have a significant effect on the depressive symptoms during pregnancy which are only partly mediated by the affective symptoms of PMS. Excessive activation of immune-inflammatory pathways may underpin the premenstrual, early and late pregnancy affective symptoms and these pathways are partly activated by ACEs. Further research should focus on the contribution of immune-inflammatory and associated nitro-oxidative pathways on the affective symptoms of PMS (better MCAS), pregnancy and the postnatal period.

## Data Availability

All data produced in the present work are contained in the manuscript

## Statements

### Author contributions

All the contributing authors have participated in the manuscript and approved the final version of the manuscript.

## Declaration of Competing Interest

The authors declare that they have no conflict of interests.

## Funding

This research is supported by the 90th Anniversary of Chulalongkorn University, Rachadapisek Sompote Fund.

## Data Access Statement

The dataset generated during and/or analyzed during the current study will be available from M.M. upon reasonable request and once the dataset has been fully exploited by the authors.

## Research involving human participants

Approval for the study was obtained from the Institutional Review Board of the University of both the Khon Kaen Hospital and Chulalongkorn University (KEF62036 and COA No.280/2019).

## Informed consent

All women gave written informed consent before participation in our study.

## Acknowledgements

We are grateful to all the participants for their time and efforts for this study. We also acknowledge with appreciation the staff of ANC at Khon Kaen hospital who greatly helped with the data collection.

## Notes

### Competing Interest Statement

The authors have declared no competing interest.

### Funding Statement

the 90th Anniversary of Chulalongkorn University Ratchadapisek Sompoch Fund, and Ratchadapisek Sompoch Endowment Fund (Grant number: RCU_H_64_034_5) and

### Author Declarations

Institutional Review Boards (IRBs) at both Khon Kaen Hospital and Chulalongkorn University (KEF62036 and COA No. 280/2019)

